# Regional comparisons of COVID reporting rates, burden, and mortality age-structure using auxiliary data sources

**DOI:** 10.1101/2021.08.18.21262248

**Authors:** Mollie M. Van Gordon, Lawrence Mwananyanda, Christopher J. Gill, Kevin A. McCarthy

## Abstract

We correct common assumptions about COVID burden and disease characteristics in high-income (HIC) versus low- and middle-income (LMIC) countries by augmenting widely-used surveillance data with auxiliary data sources. We constructed an empirically-based model of serological detection rates to quantify COVID reporting rates in national and sub-national locations. From those reporting rates, we estimated relative COVID burden, finding results that contrast with estimates based on case counts and modeling. To investigate COVID mortality by age in an LMIC context, we utilized a unique morgue study of COVID in Lusaka alongside the population attributable fraction method to account for HIV comorbidity. We calculated the comorbidity-corrected age-adjusted mortality curve in Lusaka and found it significantly skewed toward younger age groups as compared to HICs. This unexpected result recommends against the unexamined use of HIC-derived parameterizations of COVID characteristics in LMIC settings, and challenges the hypothesis of an age-structure protective factor for COVID burden in Africa. Indeed, we found overall COVID burden to be higher in Lusaka than in HICs. Concurrent with high COVID burden, many LMICs have high prevalence of other public health issues such as HIV, which compete for limited health investment resources. Given differences in age-structure, comorbidities, and healthcare delivery costs, we provide a case study comparing the cost efficacy of investment in COVID versus HIV and found that even in a high HIV prevalence setting, investment in COVID remains cost-effective. As a whole, these analyses have broad implications for interpretations of COVID burden, modeling applications, and policy decision-making.

**Significance Statement:** The analyses presented here demonstrate the power of auxiliary COVID data sources to fill information gaps, particularly for LMICs. Our results reveal differences in COVID surveillance and disease dynamics between HICs and LMICs that challenge common perceptions and assumptions about COVID in these respective contexts. We show the divergence of COVID reporting rates between HICs and LMICs and the effects on relative estimated burden. Contradicting common modeling practices, our analysis demonstrates that the age-structure of COVID mortality cannot be accurately generalized from HICs to LMICs. We find higher COVID burden in LMIC contexts than HICs particularly in younger age groups and show that investment in COVID is cost-effective even in light of other public health concerns.

---

**E**ffective counter-measures against the raging COVID-19 pandemic require accurate geographically-specific knowledge, but surveillance and data inequities present major challenges to these efforts. Surveillance systems are generally more robust in high-income countries (HICs), meaning that data from HICs tend to be more readily available and complete than data from low- and middle-income countries (LMICs) (1, 2). For lack of locally-specific data, epidemiological characteristics of COVID are often based on data from HICs, but the transferability to LMIC contexts remains an open question (3). Accounting for differences in surveillance and incorporating auxiliary data sources can help fill these data gaps and inform our understanding of COVID across contexts.

For example, official statistics on regional COVID burden are based on reported case counts (4), despite evidence of substantial case underreporting particularly in LMIC contexts (5, 6). Such differences in reporting rates can significantly alter estimates of relative disease burden across regions (7). COVID reporting rates are difficult to determine, but incorporating serological data can inform reporting rate estimates. While serology is challenging to work with, it offers some of the best information in data-sparse settings if the limitations of serological data are accounted for (8, 9).

Surveillance and reporting influence our understanding of COVID dynamics in other ways as well. For example, while there is strong evidence that COVID parameters such as infection fatality rate (IFR) vary even within HICs, data challenges in LMICs mean that HIC estimates are often used in LMIC settings (10). This practice has major implications for estimates of COVID burden and risk factors, and subsequently for policies and public health practices targeting COVID. The common understanding of IFRs and the age-structure of COVID mortality has led to hypotheses that Africa’s young population distributions have a protective effect against COVID (11), yet questions remain about impacts of comorbidity distributions, differences in disease characteristics across settings, and the role of COVID interventions in the context of other public health concerns.

We address these questions and assumptions through auxiliary data sources. Using serological modeling, we calculate reporting rates for different national and sub-national locations in HICs and LMICs. We then use that data in a reporting rate model to adjust national burden estimates accounting for differences in surveillance. Taking a closer look at a local context, we use morgue sampling data to circumvent issues introduced by sampling bias, and statistical approaches to adjust for comorbidity. With this age-disaggregated data we compare mortality age-structure and burden between LMIC and HIC contexts. Cognizant of competing public health issues, particularly in LMIC contexts, we examine relative costs and benefits of investing in COVID vaccine.

In this article, we first present our results on reporting rates across locations and relative regional COVID burden informed by serology. Next, we use auxiliary data from Zambia to examine mortality age-structure in an LMIC-specific location,and conduct an impact comparison of COVID intervention with other public health interventions. We follow with a discussion describing broader implications of our study results for programmatic and modeling applications. Finally, we describe in detail our data and methods, including serological and reporting rate modeling, mortality burden quantification, statistical correction of COVID mortality for comorbidity factors, and the cost modeling used to compare public health investments.

## Results

### Serology across World Health Organization (WHO) regions

#### Reporting rates in the European region and the region of the Americas (EURO and AMRO) are higher than elsewhere

Matching sero-prevalence estimates with reported cases in a particular location allows calculation and comparison of reporting rates(Figure 1). We take the cumulative reported cases for a location, up until the time of the serostudy, and adjust the case rate by a model of seroconversion and reversion. This case rate is lower than the raw reported cumulative case rate, as serosurveillance does not detect all infections. Reporting rates follow the dashed gray lines on the plot. In general, countries in the AMRO and EURO regions have the highest reporting rates, shown on a log scale. Note that seroprevalence cannot be directly compared across locations because of the differences in the dates of the serostudies.

**Fig. 1.**
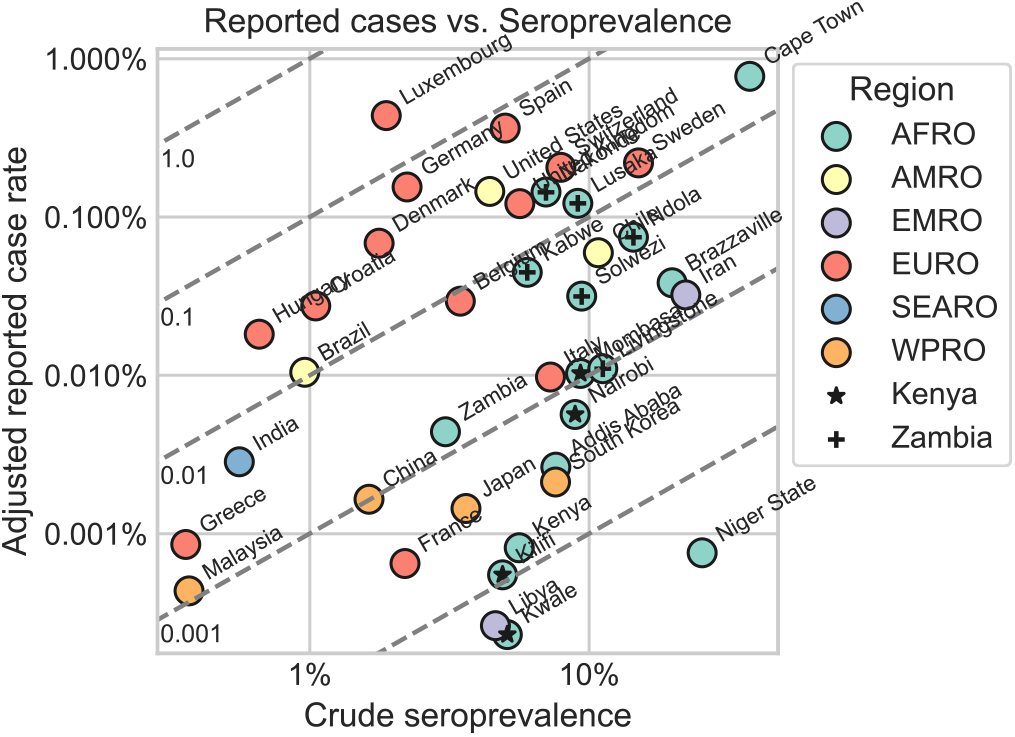
Adjusted reported cases vs. crude seroprevalence by location. Dashed gray lines indicate reporting rates, labeled on the inside of the y-axis. WHO region is shown in color; sub-national locations in Kenya and Zambia are indicated with symbols.

Because serostudy data from the African region (AFRO) are sparse, we include sub-national prevalence estimates, and match those with case data from the particular sub-national locations only. Kenya and Zambia have data for multiple sub-national locations, and in both countries reporting rates vary by more than a factor of ten. While there is a clear correlation between seroprevalence and reported cases, there are notable exceptions that appear to align with national wealth. For example, Niger State and Cape Town both have similar seroprevalence rates, but differ a thousand-fold in reported cases. South Africa is a country with considerable resources, expertise, and infrastructure for disease surveillance, and there is reason to expect that capacity to test and report COVID cases is likely better in Cape Town than across Niger State. A similar argument can be hypothesized for other low income countries on this matrix.

#### Ranked estimated infections show lowest burden in EURO

To identify the location-specific effects of reporting rates on reported case burden, we estimated infection burden across locations on the same date. Serostudies provide estimates of reporting rates for a specific window of time, but we don’t assume reporting rates are constant in time. To address this challenge, we used our point estimates of reporting rates to build an model for the relationship between reporting rate and testing rate, for which we have continuous time series data. We then used the testing rate time series to model dynamic reporting rates and unify the dates of estimated infections.

Figure 2 shows a ranking of COVID burden across locations on November 12th, 2020, the most recent date with continuous available testing data for all locations. The top plot shows the ranking based on reported cases; the bottom plot shows the ranking of estimated infections calculated using reporting rates. In the reported case burden ranking, the EURO region is high relative to other regions; however in the estimated infections ranking, EURO countries have low burden relative to other regions.

**Fig. 2.**
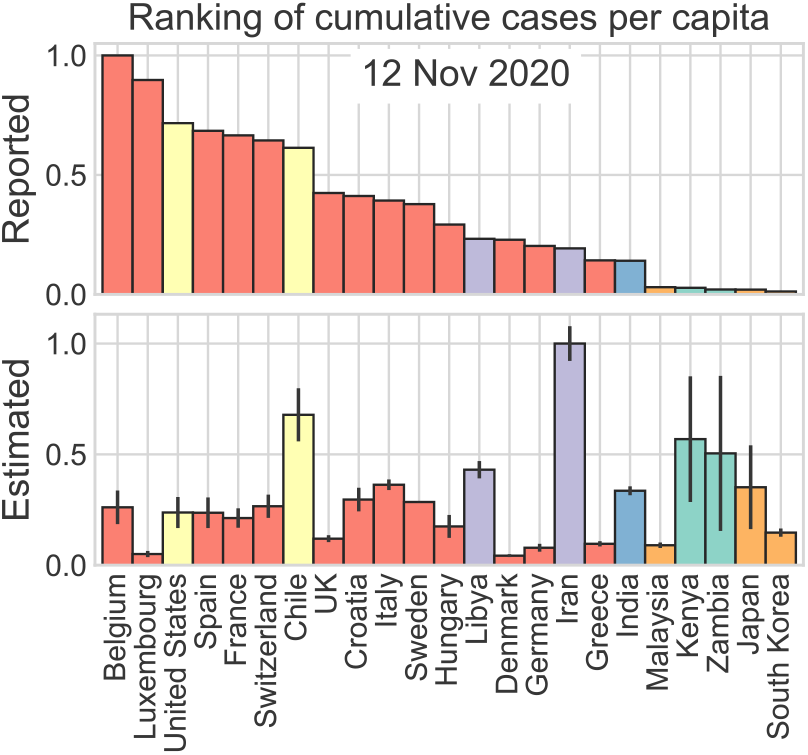
Ranking of cumulative COVID reported cases (top) and estimated infections (bottom) as of November 12th, 2020.

We note that increased estimated cases relative to reported cases occurs mainly, but not exclusively, in lower resource country settings. As with the analysis presented in Figure 1, these data support the concern that underreporting, perhaps as a consequence of resource limitations, may lead to a significant under-counting of cases in certain parts of the world.

### COVID mortality in Lusaka

#### COVID mortality burden is substantial among younger age groups in Lusaka and higher overall as compared to HICs

Based on a morgue study in Lusaka, Figure 3, top left, shows the percentage of all-cause deaths that tested positive for COVID postmortem, adjusted to exclude HIV-attributable comorbid deaths. The frequency of COVID deaths by age-bin hovers between 9% and 19% for all age groups up to age 70. COVID death frequencies in the 70s and 80+ age bins are 24% and 34%, respectively. Using age structures of population and life expectancy, COVID mortality frequencies can be translated into years of life lost (YLL) by age group, which we present for Lusaka alongside the United States for a comparison of LMIC and HIC contexts (Figure 3, top right). While as expected, YLL per capita due to COVID is highest in older age groups in the US, the pattern in Lusaka is drastically different: young age groups, instead of older age groups, suffer the highest COVID burden. In addition, total years of life lost per capita across all age-bins is higher overall in Lusaka than in the United States.

**Fig. 3.**
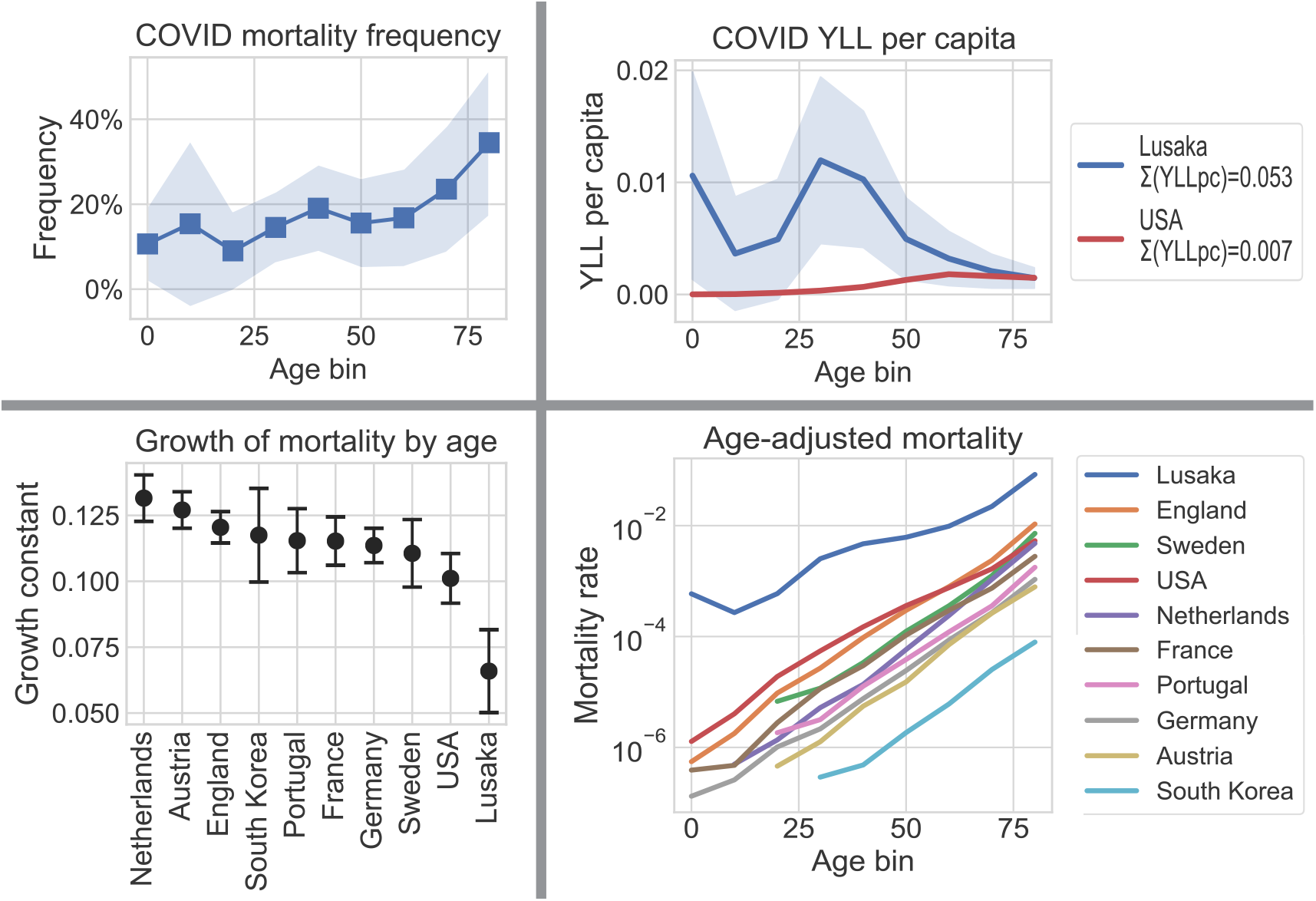
COVID mortality for Lusaka and HICs. Upper left: Frequency of COVID-positive deaths among all-cause deaths by age bin (Lusaka). Upper right: Years of life lost per capita by age bin for Lusaka and the United States. Lower right: Age-adjusted mortality for Lusaka and HICs by age bin. Lower left: Exponential rate of increase of age-adjusted mortality with age across locations. All mortality data is from early August, 2020. COVID mortality in Lusaka is adjusted to exclude HIV-attributable comorbid deaths. Shading and error bars represent 95% confidence intervals.

Plotted in Figure 3, bottom right, are age-adjusted COVID mortality rates in Lusaka vs. HICs (log scale). In all age groups, age-adjusted COVID mortality rates are higher in Lusaka than in HICs. Fitting an exponential model to each age-adjusted mortality curve provides an estimate of the rate of increase of mortality with age. The slope of this model is defined as the growth constant of the curve, plotted for multiple locations in Figure 3, bottom left. The significantly lower growth rate of mortality with age in Lusaka indicates age-adjusted mortality is skewed toward higher mortality in younger age groups as compared to HICs. While Lusaka is not necessarily representative of all LMICs, these results highlight the pitfall of generalizing COVID disease parameters from HIC to LMIC contexts.

### Costing mortality reduction

#### Investment in COVID mortality reduction should be prioritized even in the context of other public health challenges

Building on the age-specific mortality dataset from Lusaka, we used basic cost modeling to demonstrate the utility of investment in COVID mortality reduction. As HIV is the number one cause of death in Zambia, we examine HIV investment alongside COVID (12). We include for comparison the United States, a low HIV-burden HIC setting. Based on the age structures of population as well as COVID and HIV mortality, we calculate total YLL per capita for different levels of combined mortality reduction of COVID and HIV across age groups. The heat maps in Figure 4 show these results for Zambia (left) and the United States (right). Model input data are detailed in the table in Figure 4. Costing model results are shown as white isotropic lines on the heat maps, indicating cost per percent mortality reduction for COVID and HIV combined.

**Fig. 4.**
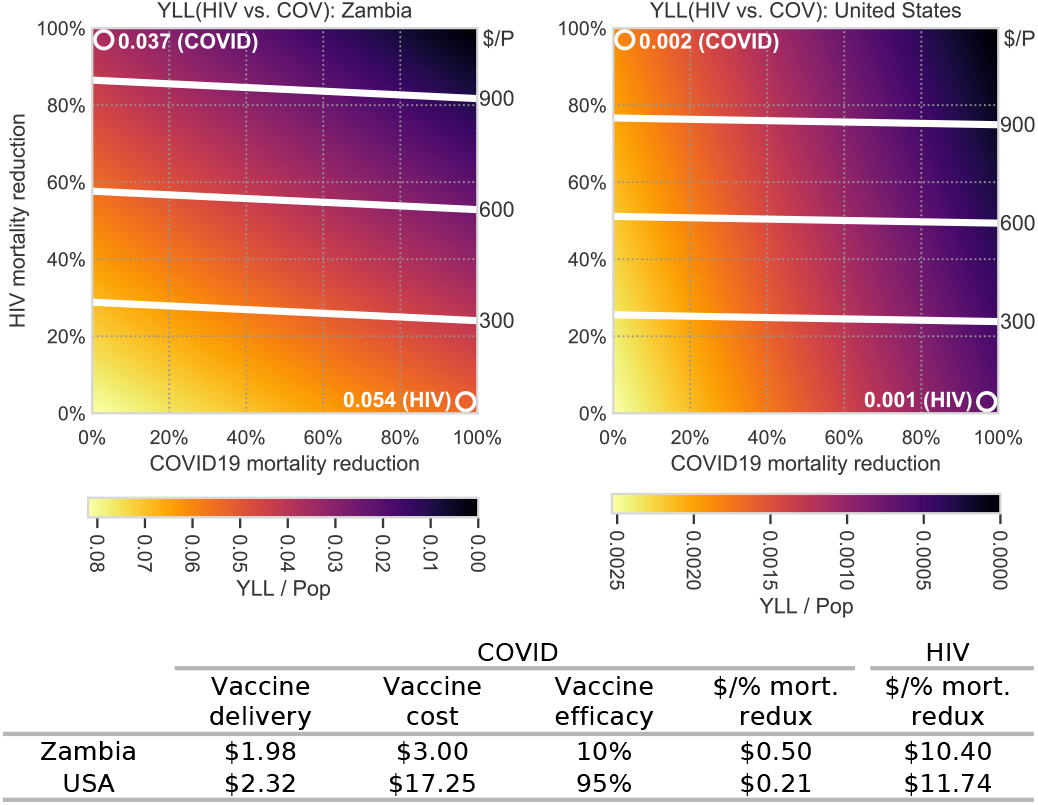
Cost models of reduction of COVID and HIV mortality for Zambia (left) and the United States (right). Axes indicate model inputs of percent mortality reduction for HIV and COVID. Heat map illustrates remaining YLL/total population for HIV and COVID combined. Cost per capita for combined HIV and COVID mortality reduction are shown in white isotropic lines, labeled on the right y-axis of each plot. White circles on the plot are annotated with the total YLL per capita for one disease only, as indicated. Table shows model inputs for COVID and HIV cost per percent mortality reduction, sources cited in the Materials and Methods section.

In both Zambia and the United States, the cost of HIV mortality reduction outstrips that of COVID mortality reduction. As a result, the gradients across the cost model isotropic lines are dominated by HIV mortality reduction in both plots. The underlying structure of YLL per capita, however, differs between the two countries. In the United States, the magnitude of COVID-attributed YLL per capita is twice that of HIV-attributed YLL per capita, as annotated by the white circles on the right plot. As a result, the gradient direction of YLL per capita in the United States is dominated by the COVID mortality reduction axis. In Zambia, HIV as the number one cause of death might be expected to dominate the gradient of YLL per capita, but COVID-attributed YLL per capita is nearly 70% that of HIV. This skews the gradient direction of total combined YLL per capita to a roughly diagonal orientation, from lower left to upper right.

The overlay of the cost/percent mortality reduction isotropic lines and the YLL per capita heat map indicate the cost efficacy of investment in HIV vs. investment in COVID. In both the United States and Zambia on a constant per capita budget, maximizing investment in COVID mortality reduction relative to HIV investment minimizes YLL per capita, the desired outcome. Note that there are many components to decision-making about public health investment, and we do not claim that our model results establish exact cost for mortality reduction or that COVID investment is definitively indicated. Rather, this analysis serves to unseat any a priori assumptions that COVID burden is insignificant in settings with other high-burden public health issues. Rather, COVID investment should be considered as a possible avenue for cost-effective reduction in total disease burden.

## Discussion

Using surveillance data auxiliary to reported COVID cases and deaths, we demonstrate that common assumptions about regional COVID burden must be reconsidered. We calculate reporting rates across locations and show the impact of regional differences on perceived COVID burden. Further, contrary to impressions derived from case counts, we establish higher burden of COVID in the African context as compared to HICs, particularly in younger age groups. This challenges predominant assumptions about the age structure of mortality rates and the protective effects of younger populations in Africa (3, 8, 13).

Combining seroprevalence data with seroconversion and reversion modeling, we calculate reporting rates across WHO regions and present estimated infections across locations. Our analysis shows high reporting rates in EURO and AMRO relative to other regions, a data-based result consistent with anecdotal understanding (1, 7). Heterogeneity of reporting rates at the sub-national level adds to the common understanding of geographic heterogeneity of COVID prevalence (14, 15). By identifying a relationship between reporting rate and testing rate, we unify the date of COVID prevalence estimates and rank countries according to relative estimated prevalence. The relatively low burden in EURO countries contradicts geographic burden distributions based on reported cases as well as modeling estimates (13, 16).

Randomly sampled morgue-based COVID testing data provides the opportunity to evaluate mortality dynamics in an African context, without the challenges associated with reporting systems. The age structure of mortality in the African setting is significantly different from HIC settings, with unexpectedly higher burden in younger age groups. In addition, overall mortality burden in the African setting outstrips that in HICs. This poses serious risks for LMIC countries where age distributions are skewed younger, directly contradicting the age-protection hypothesis. We use the mortality data alongside a simple cost model to show that even in a context with substantial other public health concerns such as HIV, COVID may be a cost-effective investment for disease burden reduction.

Our study is subject to a number of limitations, particularly as we grapple with understanding COVID dynamics in LMIC contexts. LMICs are subject to limited data availability and substantial uncertainty, which we address in part by making use of sub-national data sources including serology and COVID testing from morgue sampling. Challenges when working with serology include inconsistencies in testing protocols and sampling frameworks alongside the impacts of seroconversion and reversion on results. To address these hurdles, we focus on serostudies that do not target particular populations, and adjust estimates for seroconversion and reversion.

Necessitated by data and uncertainty limitations, some of the models we present rely on broad approximations. Modeling reporting rate as a function of testing rate, for example, is an approximation made to include countries where more detailed auxiliary data are not available. We do not attempt to estimate the magnitude of COVID burden in different locations, only their relative ranking. Finally, cost modeling is presented as a ballpark framework to evaluate COVID in the context of other public health concerns, rather than a comprehensive costing model. We use HIV as an example to compare with COVID, recognizing that there are other sources of burden and other approaches to public health investment than single disease-focused strategies. We do not attempt to model the complexities of mortality reduction dynamics, rather seeking to demonstrate that COVID should be considered as part of a public health investment portfolio.

The substantial differences in reporting rates across locations and the subsequent influence on perceived burden highlight the pitfalls of making programmatic decisions based on reported cases alone. The differences in mortality and burden between Africa and HIC contexts calls for caution when translating epidemiological age structure assumptions from HIC to LMIC contexts, and rethinking assumptions about protective factors for perceived lower burden in African contexts. Cost-effectiveness of COVID mortality reduction recommends investment even in contexts with other entrenched public health concerns and limited vaccine efficacy.

## Materials and Methods

### Data

Reporting rate analysis relies on serostudies and reported cases. In order to reduce bias in the serology data, only studies that do not target a particular population (e.g. health care workers) were included. For WHO regions other than AFRO, nationally pooled seroprevalence estimates were used (2, 17). Because serostudy data in AFRO are sparse, both national estimates and sub-national estimates were included (14, 18–22). National case rates were obtained from Our World in Data (16). Sub-national case rates were calculated from sub-national reported case counts and population data (14, 15, 19, 23–31).

Because vital reporting systems in Africa can be limited (32), we used a morgue sampling study for COVID and HIV mortality rates in Lusaka instead of reported numbers (6), all-cause mortality rate for Lusaka (32), and HIV prevalence data for Zambia (33). Zambia HIV prevalence data was extrapolated to older age bins corresponding to other data sources via a Gaussian model. For HICs, where vital reporting systems are strong, reported age-binned mortality rates for COVID and HIV were used (34, 35). Years of life lost and age-adjusted mortality require life expectancy and population by age, which were pulled from the UN World Population Prospects and the 2018 Zambia Demographic and Health Survey (36, 37). Input data for cost models included vaccine delivery, cost and efficacy for COVID, and an established costing model for HIV (38–42). Cost data from the literature were adjusted for inflation to 2021 dollars.

### Methods

#### Seroconversion and reversion

Seropositivity provides an indicator of the number of past infections, which can be compared with cumulative reported cases to estimate reporting rate. The probability of testing seropositive, however, is a function of the elapsed time from infection because of seroconversion and reversion dynamics. In other words, a serostudy does not detect all past infections. For a fair comparison of detected seropositivity with reported cases, we adjusted reported cases down to the number of those cases that could be detected by serosampling based on a model of seroconversion and reversion. We tested two models for calculating this case adjustment, each with different data requirements.

The more data-intensive model requires continuous time series of daily reported case data. The model provides an estimate of the probability for an infection to appear seropositive at time *t*, where *t* is the time between infection and the serostudy. To construct this model, we combined empirical models for the probabilities of seroconversion (43) and reversion (44), Figure S1. Where *τ* is the time between infection and seroconversion, *conv* indicates seroconversion, *rev* indicates seroreversion, *P* (*t*) indicates the probability of seroconversion or reversion at time *t*, and *p*(*t*) indicates the probability of seroconversion or reversion by time *t*, the probability of an infection being seropositive at time *t* is as follows:

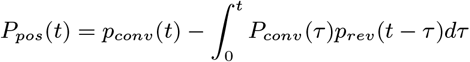

Because the empirical models from the literature include only about four months of data, we used the last month of the combined empirical model to extrapolate out to a year using a log-linear regression. The full *P*_*pos*_ model is shown in Figure S2.

The calculation for a point estimate of reporting rate using this model of seropositivity is then as follows, where *T*_*k*_ is the date the serostudy *k* was conducted, *R* is reporting rate, *c* is daily reported cases, and *S* is seropositivity rate:

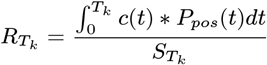

While continuous daily case reporting data largely exists for HICs, LMICs do not necessarily report data at a daily frequency, particularly for sub-national locations. In order to be able to include more LMIC serology studies, we also developed a less data-intensive model for the case count adjustment to approximate seroconversion and reversion dynamics. In this model, only two data points for cumulative cases are required: cumulative cases at 21 days and 60 days before the serostudy. These time delays were selected heuristically to account for seroconversion and reversion, respectively. Only cases reported within these time bounds are then used for the reporting estimation. Where *C* is cumulative cases:

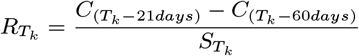

We compared results from the two different models for adjusting cases and estimating reporting rate. We found the less data-intensive model to be a good approximation of the more complete probability-based model, see Figure S3. We used the second, less data-intensive model for Figure 1 to include sub-national locations, and the probability-based model for Figure 2.

#### Dynamic reporting rate modeling

With reporting rate estimates based on serostudies each conducted at a different time *T*_*k*_, it remains to unify dates of estimated infection rates for comparison across locations. To allow reporting rates to vary over time, we constructed a hybrid reporting rate model based on the reporting rate at the time of a serostudy and the log-log relationship between testing rate and reporting rate in the serostudy locations.

As the serostudies we used are relatively early in the pandemic, we approximated reporting rate up until the time of a serostudy as the reporting rate at the time of the serostudy. This offsets under-estimation of reporting rates early in the pandemic when testing policies were largely symptomatic and testing rates were low. For dates after the serostudy, we allowed the reporting rate to vary with testing rate. The parameters of the relationship between these two variables were established by a log-log regression, illustrated in Figure S4. Where *α* and *β* are parameters of the regression fit and *E* is testing rate:

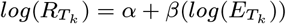

Figure S5 illustrates output from the hybrid reporting rate model for a few example countries. The modeled reporting rate over time was then used to calculate cumulative estimated infections per capita at time *T* :

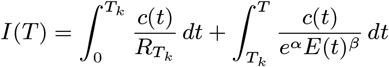

#### Mortality and years of life lost

Formulations for mortality and years of life lost in Lusaka were determined by available data. Despite the challenges that the number of deaths in Lusaka at large is difficult to determine and the catchment of the Lusaka morgue study is unknown, the randomly sampled morgue data made it possible to create formulations for age-adjusted mortality and YLL that use only available information.

By definition, where *pop*(*age*) is population by age bin; *Pop* is total population; *deaths*(*age*) is deaths by age bin; *Deaths* is total deaths; and *e*(*age*) is life expectancy by age:

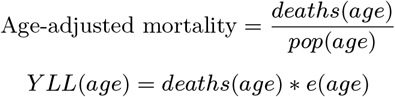

Where *morgue*(*age*) indicates the number of deaths by age bin from the morgue sample, *Morgue*_*AC*_ indicates the total all-cause deaths in the morgue sample, and *Deaths*_*AC*_ indicates the total all-cause deaths in Lusaka, we approximated 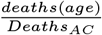 as 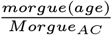. 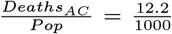 For We used the crude death rate for Lusaka from (32): 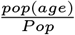, we used the urban population distribution from (37). Age-adjusted mortality then becomes:

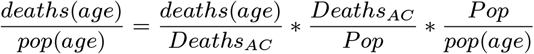

YLL is calculated per capita:

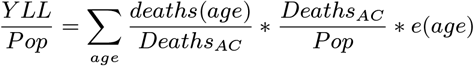

See Figure S6 for a breakdown of the frequencies of COVID and HIV within age-binned deaths. The methodology for the calculations of the increase of mortality rates with age are based on Goldstein & Lee (45).

#### Population attributable fraction

For deaths that are both COVID+ and HIV+, we attributed some to COVID and some to HIV based on the population attributable fraction method (46). Where *deaths*_(COV+)_ denotes all COVID+ deaths, *deaths*_(COV+|HIV+)_ denotes COVID+ deaths that were also HIV+, *deaths*_(COV+|HIV-)_ denotes COVID+ deaths with a negative or undetermined HIV status, and *HIV* indicates HIV prevalence, the population attributable fraction, calculated by age bin, is formulated as follows:

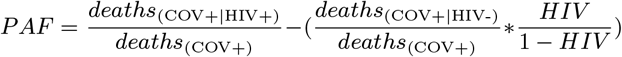

And deaths attributed to COVID by age bin are as follows:

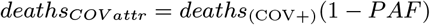

#### Cost modeling

Cost per percent mortality reduction for COVID was formulated as follows:

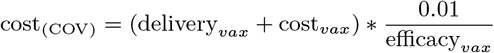

Existing literature informed vaccine delivery cost estimates (38). Vaccine cost and efficacy was calculated for the United States based on mRNA vaccines against wild type COVID (39, 40). For Zambia, vaccine cost and efficacy was calculated based on AstraZeneca against variant B1.351 (39, 41). Cost per percent mortality reduction for HIV came directly from the literature (42).

## Data Availability

Data used in this manuscript are public except data from Mwananyanda et al. 2021.

## ACKNOWLEDGMENTS

We gratefully acknowledge the team responsible for the collection and processing of the morgue data from Zambia, namely, William MacLeod, Geoffrey Kwenda, Rachel Pieciak, Zachariah Mupila, Rotem Lapidot, Francis Mupeta, Leah Forman, Luunga Ziko, Lauren Etter, and Donald Thea (6). Thanks also to Mandy Izzo for her editorial support.

