## Supplemental Information for "Regional comparisons of COVID reporting rates, burden, and mortality age-structure using auxiliary data sources"

### This PDF file includes:

Figs. S1 to S6 (not allowed for Brief Reports)
SI References

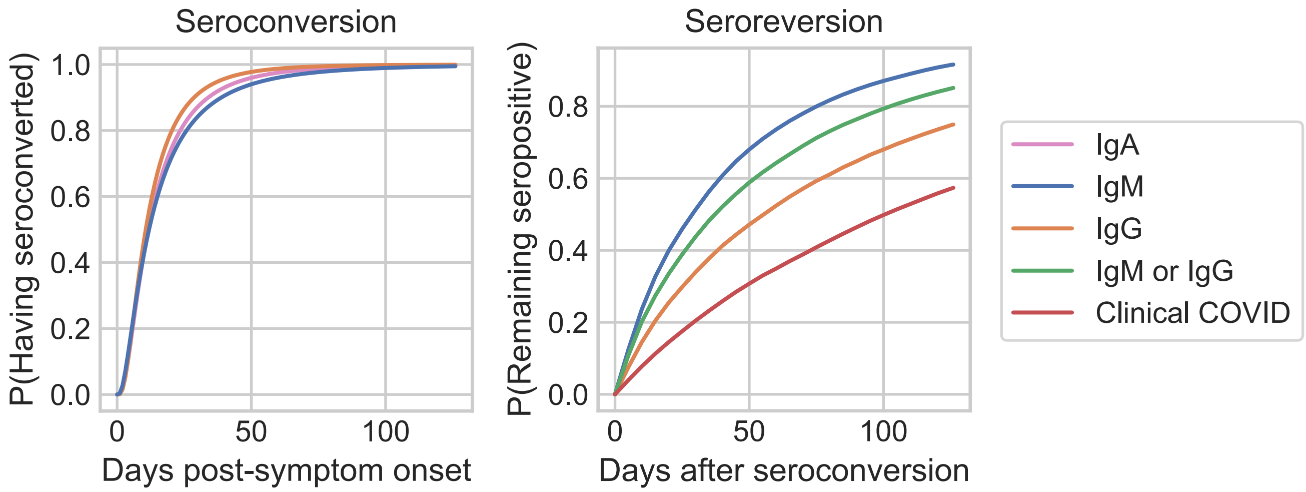


**Fig. S1.** Empirical data-based models for seroconversion (left) and seroreversion (right), reproduced from (1, 2), respectively. To align with typical isotype and sampling practices for testing, IgG is selected for seroconversion and clinical COVID is selected for seroreversion to combine into the empirical model plotted as part of Figure S2.

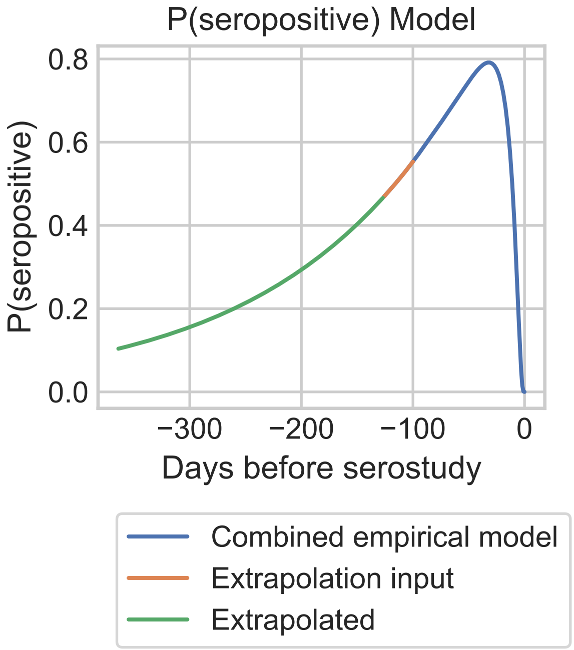


**Fig. S2.** Model for probability of being seropositive at the time of a serostudy. X-axis represents the days before a serostudy on which infection occurred. The empirical seropositivity model is constructed from a combination of the models shown in Figure S1 and extrapolation as described in the Methods section of the main text.

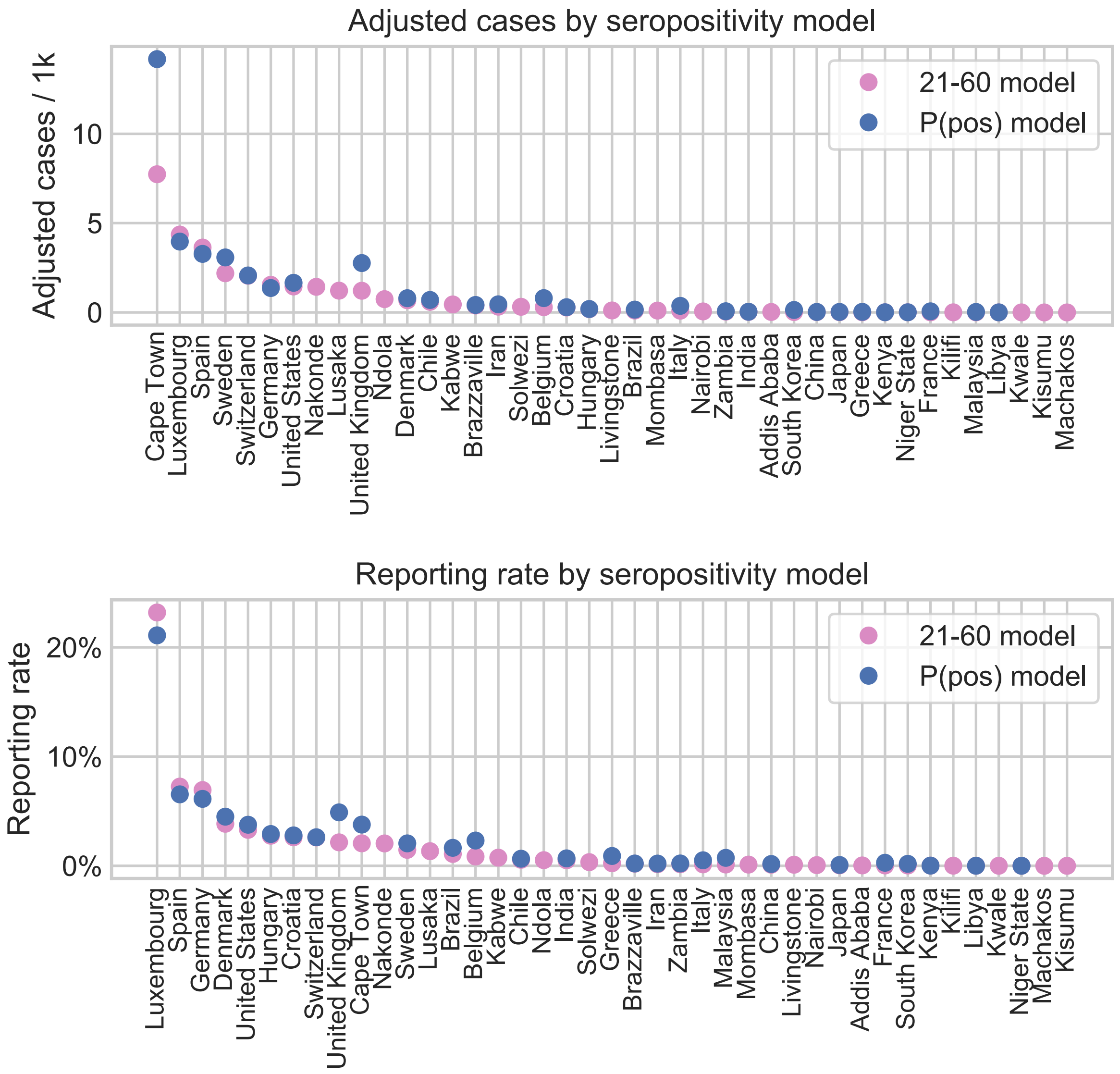


**Fig. S3.** Comparison of results from the two different models used for adjusting expected cases at the time of a serostudy. Pink shows the less data-intensive model; blue indicates the more data-intensive model. Top panel shows adjusted cases; bottom panel shows calculated reporting rate. Data are shown for the date of serostudy $(T_k)$ at each location. Each panel is sorted by the values from the less data-intensive model; the differences between pink and blue in each panel are relevant rather than the order of the locations presented. Some sub-national locations do not have estimates for the P(pos) model due to data limitations.

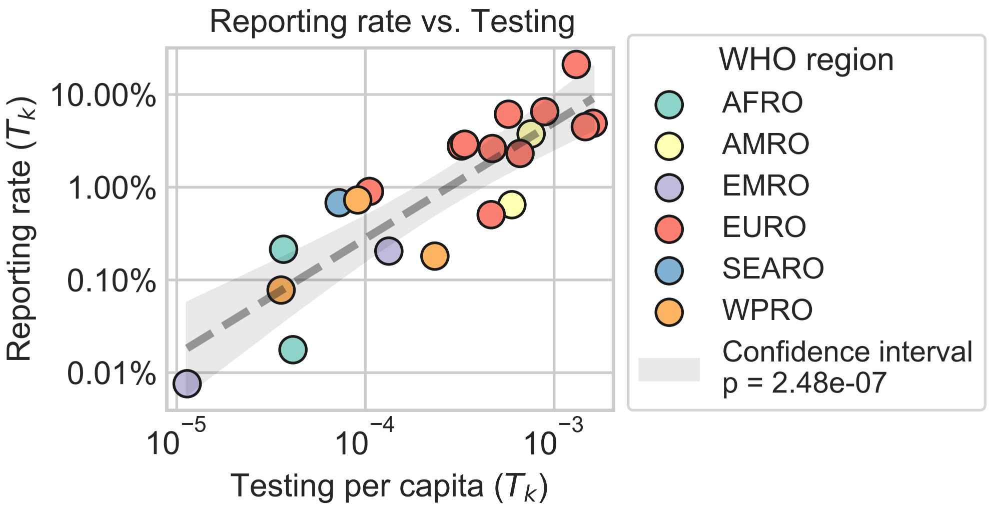


**Fig. S4.** Log-log relationship between testing rate and reporting rate used for constructing the dynamic reporting rate model. Points are color-coded according to WHO region. Data plotted are snapshots of the testing rate and reporting rate at the time of that location's serostudy $(T_k)$. $T_k$ reporting rates are estimated using the P(pos) model. Regression fit is shown in gray with a 95% confidence interval. P-value of the fit is 2.48e-07.

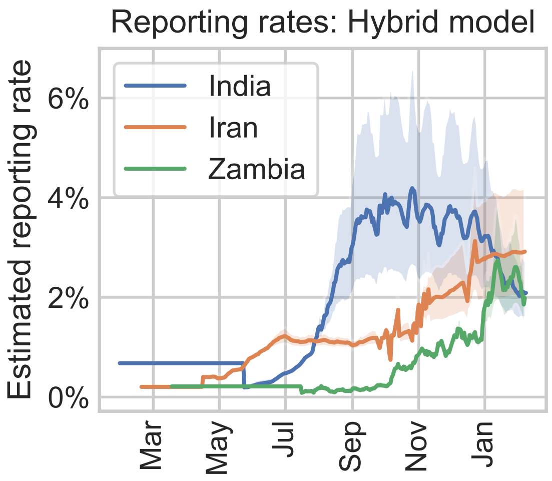


**Fig. S5.** Estimating reporting rates over time for three sample countries based on the hybrid reporting rate model. 95% confidence intervals shown with shading. Discontinuities are observed at the serostudy date for each country as a result of the transition between the two models used to make the hybrid reporting rate model: the static reporting rate calculated at the time of that country's serostudy and the dynamic reporting rate model.

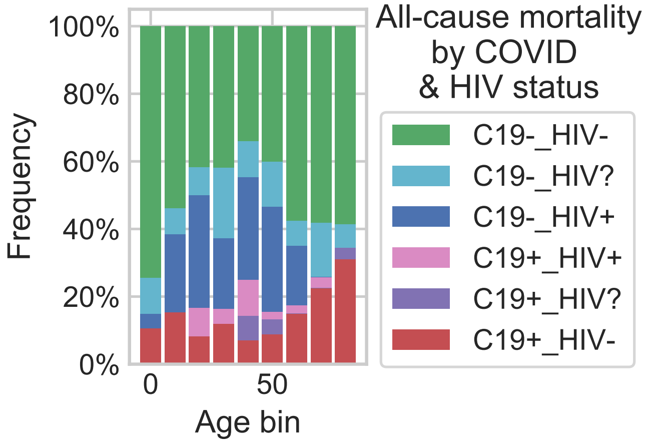


**Fig. S6.** All-cause mortality frequency within age groups by COVID and HIV status. "HIV?" indicates unknown HIV status.
